# A new compartment model of COVID-19 transmission: The broken-link model

**DOI:** 10.1101/2022.03.04.22271940

**Authors:** Yoichi Ikeda, Kenji Sasaki, Takashi Nakano

**Author notes:** Correspondence: e-mail^1^; e-mail^2^.

## Abstract

We propose a new compartment model of COVID-19 spread, the broken-link model, which includes the effect from unconnected infectious links of the transmission. The traditional SIR-type epidemic models are widely used to analyze the spread status, and the models show the exponential growth of the number of infected people. However, even in the early stage of the spread, it is proven by the actual data that the exponential growth did not occur all over the world. We consider this is caused by the suppression of secondary and higher transmissions of COVID-19. We find that the proposed broken-link model quantitatively describes the mechanism of this suppression and is consistent with the actual data.

## 1. Introduction

Since the first patient of novel coronavirus infectious disease (COVID-19) was reported in Wuhan, China, COVID-19 has spread all over the world. In order to save the lives from the threat of COVID-19 and maintain social activities from the viewpoint of economy, it is vital to ascertain accurately the status of the spread.

The SIR (susceptible-infected-removed) model and its family such as the SEIR (susceptible-exposed-infected-removed) model have been widely used compartment models trying to describe the projection of COVID-19 spread. The SIR model was first applied to the plague in the island of Bombay over the period Dec. 1905 to July 1906 [1]. The first order coupling between susceptible and infected people was assumed, and such treatment was justified for the plague mediated by carrier rats which form a mean field of the plague, and thus the susceptible people have an equal probability of being infected. Indeed, the calculated epidemic curve during the period of epidemic roughly agreed with the reported numbers. One of typical features of the SIR-type models is that the models predict the exponential growth of the number of infected and removed people for the early stage of the spread [2].

Meanwhile, the indicator of the spread rate, what is called the K-value, defined by *K*(*t*) = 1 − *R* (*t* − 7)/*R* (*t*) with *R*(*t*) being the cumulative number of confirmed cases at day *t* from a reference date, exhibits nonexponential growth of *R*(*t*) even in the early stage of the spread but exhibits approximate linear decrease of the K-value transition universally in many countries [3]. The linearly decreasing behavior of the K-value transition was well reproduced by the phenomenologically developed constant attenuation model [3], where *R*(*t*) is expressed as *R*(*t*) = *R*(0) exp(*a*(*t*) *t*), and *a*(*t*) is defined by the geometric progression, *a*(*t*) = exp[−(1 − *k*)]*a*(*t* − 1) with a constant attenuation factor *k*. Based on the constant attenuation model, it was found that *R*(*t*) follows the Gompertz curve [4-6].

In this paper, we propose a new compartment model, the *broken-link model*, in order to microscopically understand why the COVID-19 transmission follows the Gompertz curve. The model is naturally derived from the observation of suppression of COVID-19 transmission in the secondary cases generated by the primary ones [7]. We also apply the model to the epidemic surges generated by Delta (*δ*) and Omicron (*o*) variants in Japan, South Africa, Unites States, France and Denmark.

## 2. Materials and Methods

To derive the broken-link model, we start with the SIR model. In the SIR model, we partition the total population into three compartments: susceptible, infected and removed individuals, and represent the numbers of three compartments at time *t* by *S*(*t*), *I*(*t*) and *R*(*t*). The SIR model is then described as coupled ordinary differential equations (ODEs),

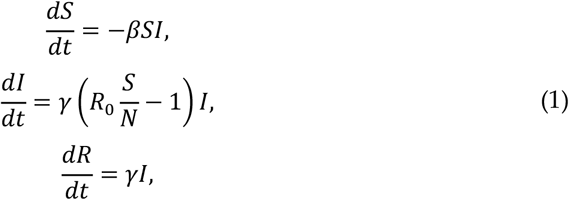

where *β* and *γ* are contact and removal rates of infections, respectively. The basic reproduction number is denoted by *R*_0_ = *βN*/*γ* with *N* being the total population number. When the cumulative number of infected persons *R*(*t*) is much less than the total population, *S*(*t*) can be approximated by *N*. Then one finds the exponential growth of *I*(*t*) and *R*(*t*) which cannot be inevitable unless the contact and removal rates are assumed to be constant in the period of epidemic.

One of good indicators to find out the behavior of COVID-19 transmission is the K-value. The analysis using the K-value has revealed that the cumulative number of confirmed cases *R*(*t*) follows the Gompertz curve even in the early stage of the spread, where the herd immunity has not been achieved at all. A natural reason is that COVID-19 spread through the contact and/or local droplet processes. As reported in Ref. [7], the secondary transmission generated from the primary cases in non-close environments is highly suppressed.

We model the suppression of the secondary and higher transmission in terms of compartment models. According to Ref. [7], all the transmission links are not connected to the next generation. When the links are connected through the probability *k*, in other words through the broken-link probability (1 − *k*), the subsequent transmissions are not generated. Therefore, we cut these contributions from a transmission tree as shown in Figure 1 (a).

**Figure 1.**
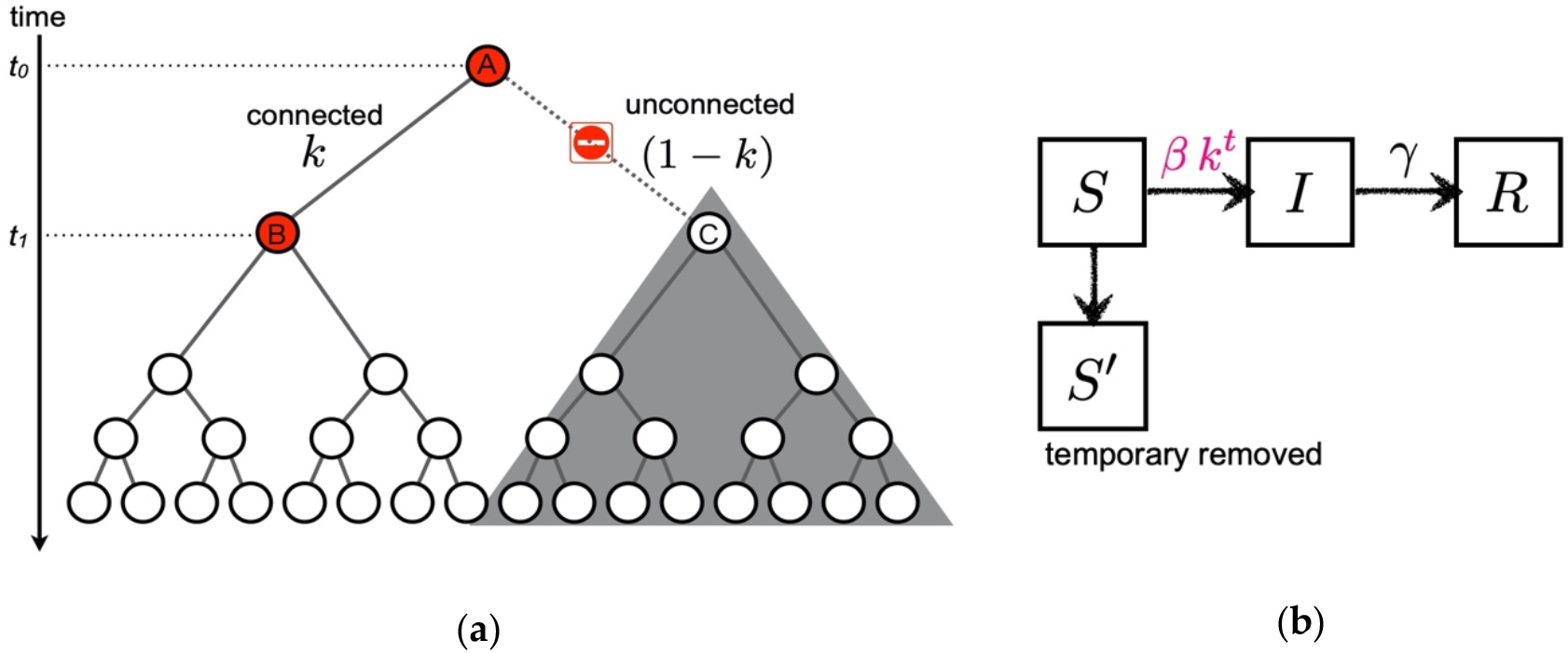
The idea to formulate the broken-link model. (**a**) Cartoon of the suppression of secondary infection in a transmission tree (*k* = 1/2 case). When the transmission link from the primary infected individual A to the secondary candidate C is unconnected at time *t*_1_, subsequent transmissions starting from C are not generated as denoted by the shaded area. (**b**) Compartments of the broken-link model. The temporary removed compartment *S*′ is introduced due to the suppression of the secondary and higher transmissions. Contrary to the SIR model, the coupling between susceptible *S* and infected *I* becomes time dependent in the broken-link model.

Now, we formulate the *broken-link model*. In addition to the *S, I* and *R* compartments, it is natural to introduce the *S*′ (temporary removed) compartment due to unconnected transmission links as shown in Figure 1 (b). The time evolutions of *S*(*t*), *I*(*t*), and *R*(*t*) are respectively expressed by the following coupled ODEs:

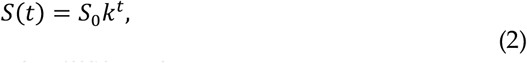

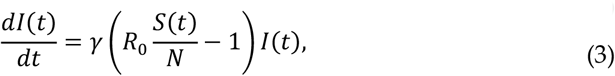

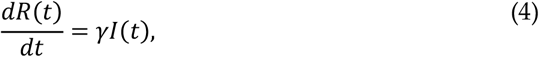

where *S*_0_ in Eq. (2) is the number of susceptible people who are potentially under the threat of transmissions in each epidemic wave, and the number of temporary removed people is given by *S*^′^(*t*) = *S*_0_(1 − *k*^*t*^). It is worth mentioning that we neglect the tiny contribution to decrease in *S* through the contact term −*βSI* in Eq. (1), which gives less than 1% contribution, since the number of infected people is two or three orders of magnitude smaller than the total population.

The analytic solutions of *I*(*t*) and *R*(*t*) are found as

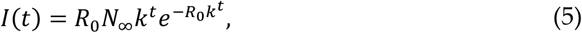

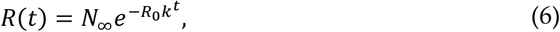

with *γ* = − ln *k* in Eqs. (3) and (4), and *N*_∞_ = *R*(0) exp(*R*_0_) which represents the cumulative number of infected people in each infection wave generated by a coronavirus with the basic reproduction number *R*_0_. In Eq. (6), we can see that the cumulative number *R*(*t*) satisfies the Gompertz curve. It also turns out that the probability *k* is equivalent to the constant attenuation factor [3], so that the phenomenologically introduced constant attenuation is consistent with the suppression of transmissions due to the unconnected transmission links.

There are two remarkable findings based on the broken-link model. First, the shapes of epicurves of the daily and cumulative confirmed cases are governed by the value of the probability *k*, and the magnitudes of the cases are proportional to exp(*R*_0_) in each infection wave. Second, the basic reproduction number *R*_0_ is inversely proportional to − In *k* ≅ (1 − *k*), namely inversely proportional to the broken-link probability when *k* is close to one. Therefore, even though only the small regional difference in the probability *k* is obtained from the actual data, the orders of magnitudes of the confirmed cases can be largely different.

## 3. Results

The surges of COVID-19 occurred in various countries. We investigate the structure of each surge of COVID-19 assuming the broken-link model. All the data are taken from COVID-19 Data Repository by the Center for Systems Science and Engineering (CSSE) at Johns Hopkins University [8]. In the analysis, the bump structure appears in daily confirmed cases as a counterpart of the Gompertz function in the cumulative number. Such bump is called a *wave* in this article. The constant trend in daily confirmed cases is described as a *baseline* which corresponds to the endemic spread and yields the linear trend in the cumulative number.

### 3.1. The δ epidemic surge in Japan

We first look at the surge caused by the *δ* variant in Japan from late June to the end of September 2021. The epicurve in Figure. 2 (a) is decomposed into three partial waves and a baseline component. Such decomposition is validated from the behavior of the K-value transition in Figure 2 (b), because there are three bumps after quick reduction as 1/*t*, which indicates the existence of a baseline in daily confirmed cases. Based on this fact, the cumulative number is fitted by three Gompertz curves and a baseline. The results reproduce both the number of daily confirmed cases and the K-value very well. The result of the fit parameters for each Gompertz curve in the *δ* surge is summarized in Table 1.

**Figure 2.**
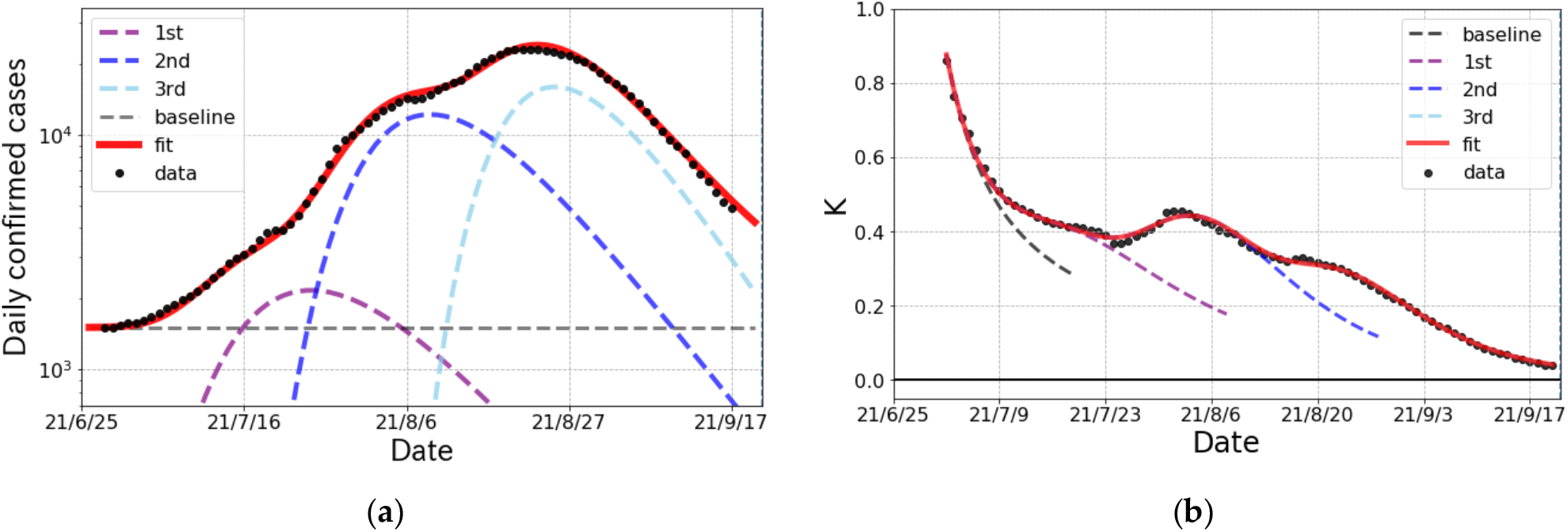
The epicurve of the *δ* surge of COVID-19 spread in Japan from June to September 2021. (a) Logarithmic plot of the number of daily confirmed cases (one week average) and fit result (solid curve). The fit was performed with three partial waves and a baseline denoted by dashed lines. (b) The observed data and fit result of the K-value.

**Table 1.**
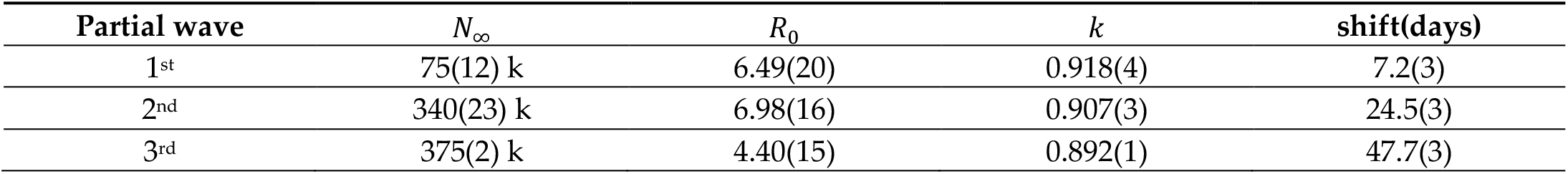
The parameters of the Gompertz curves in the *δ* surge in Japan. The *N*_∞_, *R*_0_ and *k* are the cumulative number of infected people, basic reproduction number and connected probability of transmission links, respectively. The “shift” stands for the onset of a partial wave from the reference date (06/25 2021). The statistical errors evaluated by the jackknife method are represented in the parentheses.

| Partial wave | $N_\infty$ | $R_0$ | $k$ | shift(days) |
| --- | --- | --- | --- | --- |
| 1<sup>st</sup> | 75(12) k | 6.49(20) | 0.918(4) | 7.2(3) |
| 2<sup>nd</sup> | 340(23) k | 6.98(16) | 0.907(3) | 24.5(3) |
| 3<sup>rd</sup> | 375(2) k | 4.40(15) | 0.892(1) | 47.7(3) |

### 3.2. The o epidemic surge in Japan

The trend in confirmed cases and the K-value for the *o* surge in Japan is shown in Figure 3. The first infected person with *o* variant as a community-acquired infection was reported on 22^nd^ December 2021 in Osaka prefecture and then *o* variant spiked nationwide. As seen in Figure 3, the decomposition of the *o* surge into two partial waves was justified by the trend of the K-value.

**Figure 3.**
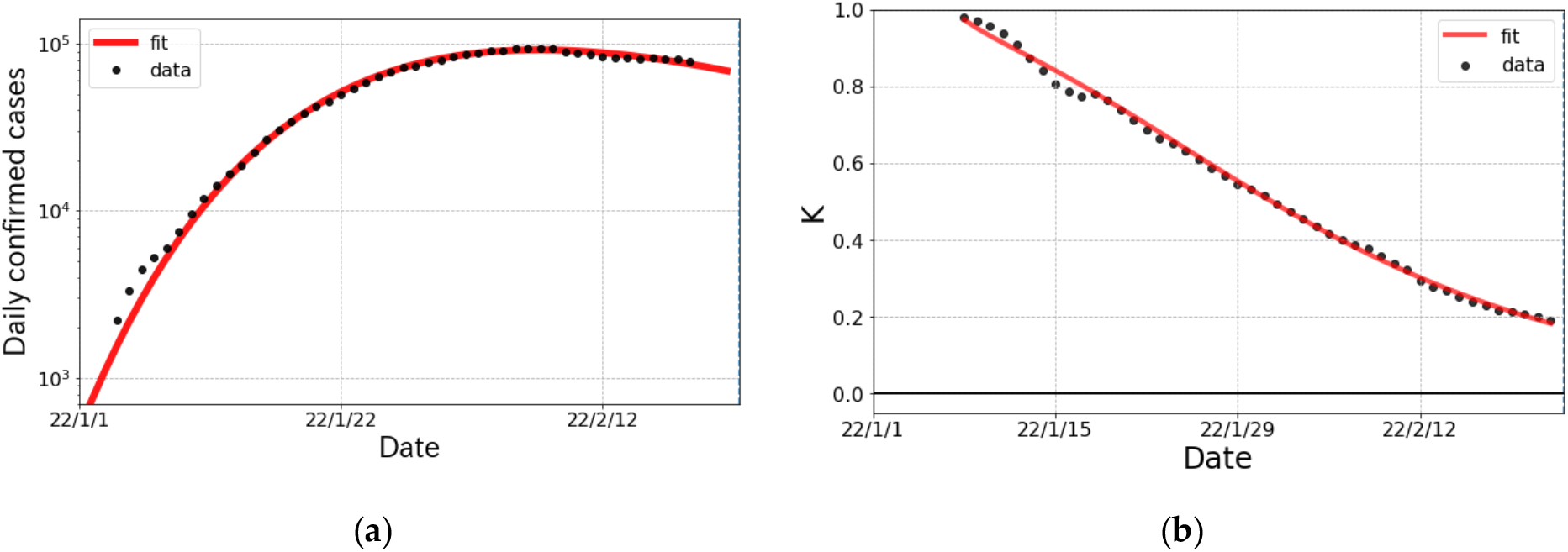
The *o* surge in Japan from January to February 2022. (a) Logarithmic plot of daily confirmed cases (one week average) and fit result. The fit was performed with a single partial wave. (b) The observed data and fit result for the K-value.

The fit parameters of the Gompertz curves for the *o* surge in Japan are summarized in Table 2. The broken-link probability (1 − *k*) is smaller than that in the *δ* surge, which implies that the subsidence of the *o* surge gets slow comparing to the *δ* surge. The cumulative number of confirmed *o* cases in Japan is predicted to be about 5 times larger than that of the *δ* case.

**Table 2.** The parameters of the Gompertz curve for the *o* surge in Japan. The definition of the parameters is the same as in Table 1, but the reference date is 1^st^ January 2022.

| Partial wave | $N_{\infty}$ | $R_0$ | $k$ | shift(days) |
| --- | --- | --- | --- | --- |
| 1<sup>st</sup> | 4,332(6) k | 10.4(1) | 0.944(1) | -4.3(1) |

### 3.3. The status of the δ and o surges in other countries

In this subsection, we survey the status of the *δ* and *o* surges in the other countries. In South Africa, where the *o* variant was reported in the world for the first time, the surge emerged on 23^rd^ November 2021 shown in Figure 4 (a). The wave in South Africa passed a peak at mid-December 2021. In U.S., the *o* surge started rising at beginning of December 2021 with huge infectivity as shown in Figure 5 (b). In France shown in Figure 5 (c), two partial waves were confirmed in the epicurve and the magnitude of 2^nd^ wave was much larger than that of 1^st^ one. In the case of Denmark, where different kind of *o* variant was reported to spread, we were able to confirm the existence of three partial waves in the surge from November 2021 to mid-February 2022 in Figure 5 (d). Again, we easily see that the number of daily cases at peak was getting larger and larger than each prior wave. The results of the fit parameters are summarized in Table 3.

**Figure 4.**
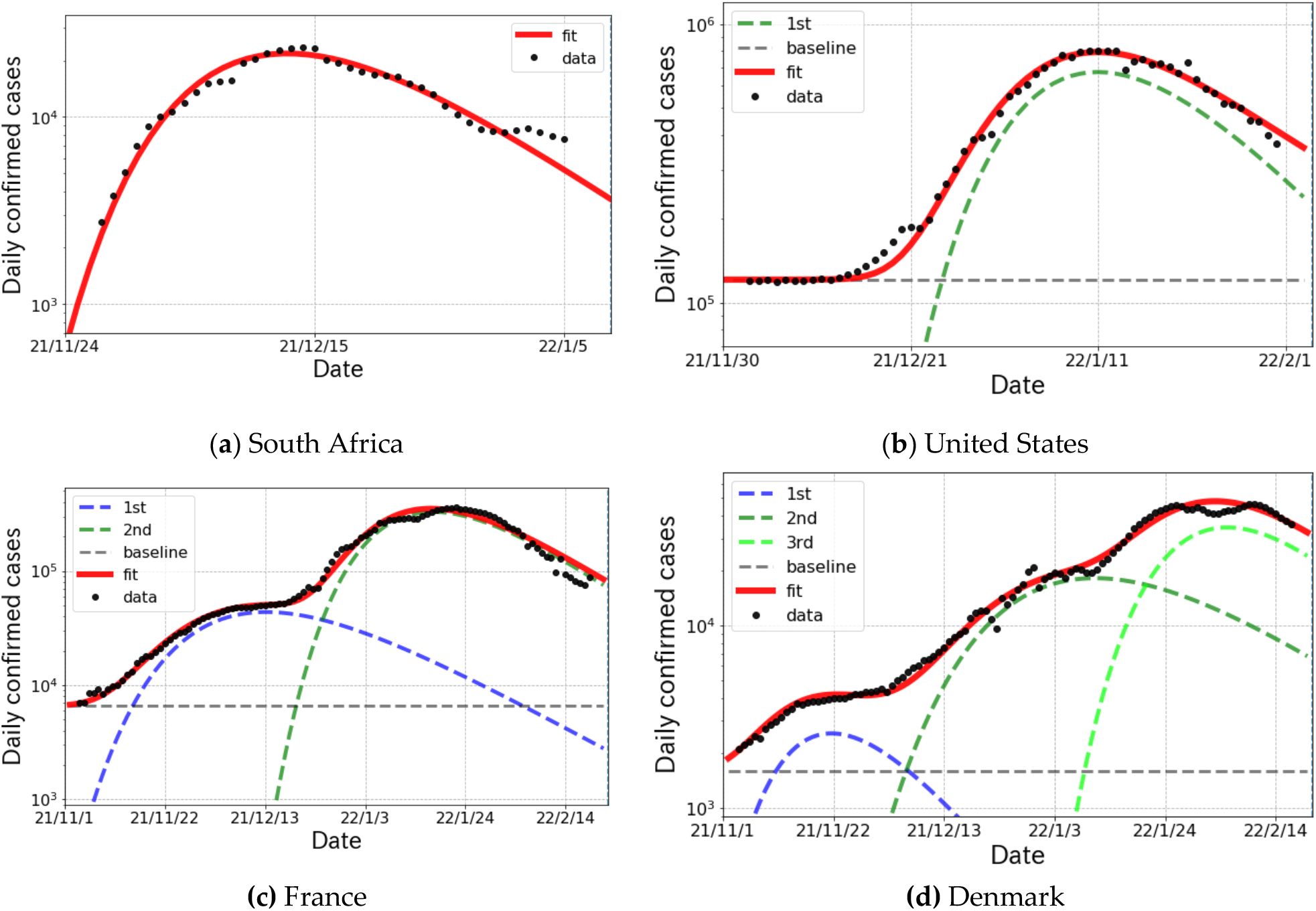
Logarithmic plot of daily confirmed cases (one week average) and fit results for *δ* and *o* surge in South Africa, United States of America, France and Denmark from November 2021 to February 2022.

**Figure 5.**
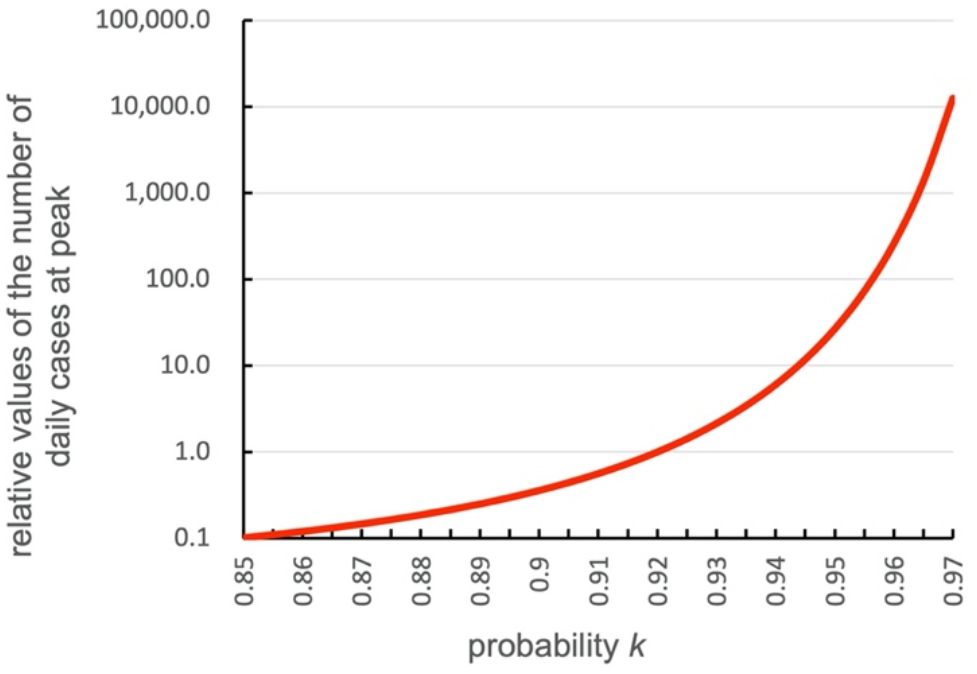
The predicted ***k*** dependence of the relative values of the number of daily confirmed cases at peak positions in the logarithmic scale. The relative value is normalized at ***k*** = **0. 92**, which was obtained in the 1^st^ epidemic surge in April 2020, Japan [3]. The value is proportional to −(**In *k***) **exp**(−***a*/ In *k***) in the model. The case with ***a*** = **0. 5** is shown.

**Table 3.** The parameters of the Gompertz curves for other countries from November 2021 to February 2022. The definition of the parameters is the same as in Table 1, but the reference dates are 11/24, 11/30, 11/01 and 11/01 for South Africa, United States, France and Denmark, respectively.

| Region | Partial wave | $N_{\infty}$ | $R_0$ | $k$ | shift(days) |
| --- | --- | --- | --- | --- | --- |
| South Africa | 1<sup>st</sup> | 592(1) k | 8.98(11) | 0.905(1) | -3.7(2) |
| United States | 1<sup>st</sup> | 22,411(269) k | 9.68(6) | 0.922(1) | 13.6(4) |
| France | 1<sup>st</sup> | 2,233(235) k | 7.18(22) | 0.949(2) | 4.3(6) |
|  | 2<sup>nd</sup> | 13,330(16) k | 9.92(1) | 0.935(1) | 42.4(1) |
| Denmark | 1<sup>st</sup> | 84(1) k | 6.16(64) | 0.924(1) | -3.0(6) |
|  | 2<sup>nd</sup> | 1094(124) k | 9.93(28) | 0.956(2) | 19.4(1.0) |
|  | 3<sup>rd</sup> | 1,401(6) k | 8.00(25) | 0.937(1) | 63.3(5) |

## 4. Discussion

As a new compartment model of COVID-19, we have proposed the broken-link model, where the suppression of secondary and higher transmissions is taken into account. The model predicts the Gompertz curve for the cumulative number of confirmed cases, which is consistent with the observations shown in Figures 2 to 4. In the model, the shape of epicurves is controlled by the probability *k*, and the magnitude is proportional to exp(*R*_0_) in which the basic reproduction number *R*_0_ is obtained as *R*_0_ = −*a*/In *k* ≅ *a*/(1 − *k*) for *k* ≅ 1 with a constant *a*. Therefore, the small regional difference of the probability *k* observed in Section 3 is enhanced in the numbers of daily and total confirmed cases. Shown in Figure 5 is the predicted *k* dependence of the number of the daily cases at peak in the model.

From Ref. [3], the mean value of the probability *k* in Japan was found to be *k* = 0.92 (8% for the broken-link probability), which is also consistent with the 1^st^ partial waves in the *δ* surge as shown in Table 1. On the other hand, for example in France, the broken-ink probability was approximately 30% smaller as shown in Table 3. This difference gives about 12 and 17 times larger in the number of daily cases at a peak and the cumulative number than those in Japan, respectively. The regional *k* difference would be attributed to the immune response to coronaviruses [9]. Indeed, due to double vaccination, about 20% and 30% increases in the broken-link probability were observed for the 2^nd^ and 3^rd^ *δ* partial waves in Japan, respectively. Thanks to the double vaccination, the 2^nd^ and 3^rd^ partial wave were suppressed to about 40% and 30%, respectively.

It is notable that the onset of epidemic surges or even partial waves has synchronized the appearance of new variants of coronaviruses in country to country. In Japan, the genomic surveillance by NIID (The National Institute of Infectious Diseases) [10] testified that the *δ* surge in Figure 2 was caused mainly by AY.29 and following AY.29.1 in terms of PANGO (Phylogenetic Assignment of Named Global Outbreak) Lineages [11]. It is interpreted that the temporary removed susceptible people from the transmission links of the *α* variant, which was already spread nationwide before the *δ* variant emerged, were brought back under the threat of the *δ* surge. The emergence of a new partial wave was also attributed to the appearance of new variants having a stronger transmissibility than the others.

It is also important to investigate the situation in other countries with respect to the genomic surveillance. In South Africa shown in Figure 5 (a), BA.1 cases were dominant during the *o* surge and then BA.2 cases gradually increased from mid December 2021. In U.S. as shown in Figure 5 (b), the *o* surge was caused by BA.1 and BA.1.1 of which transmissibility rates are expected to be similar. Thus, the fit with single Gompertz curve worked very well. For the case of France shown in Figure 5 (c), the 1^st^ and 2^nd^ waves were caused by the *δ* and *o* variants, respectively. This fact was able to be confirmed by the results of *R*_0_ given in Table 3, because the *R*_0_s in the 1^st^ and 2^nd^ waves are consistent with the typical values for the *δ* [12] and *o* [13] variants, respectively.

The situation is slightly complicate in Denmark shown in Figure 5 (d). The 1^st^ wave was generated by the *δ* variant and the others were caused by the *o* variant. The genomic surveillance report from the *outbreak.info* [14] indicated that BA.2 cases emerged from mid-December 2021 and became dominant at the end of January 2022. According to the report [14], we find two points that the 2^nd^ and 3^rd^ waves were caused respectively by BA.1 and BA.2, and BA.2 has enough strong infectivity to generate a new wave in daily confirmed cases.

## 5. Conclusions

We proposed a new compartment model of COVID-19 spread, the broken-link model, which includes the effect from unconnected infectious links of the transmission. The model took into account the suppression of secondary and higher transmissions of COVID-19. The cumulative number of confirmed cases *R*(*t*) in the model satisfies the Gompertz curve whose parameters are characterized as the cumulative number of infected people *N*_∞_, the basic reproduction number *R*_0_ and the connection probability of transmission links *k*, which was defined as the attenuation factor in the previous paper [3].

The model applied to the actual data for epidemic surges of coronaviruses in Japan, South Africa, Unites States, France and Denmark. From these results with the detailed genomic surveillance, we found that the onset of a partial wave has synchronized the appearance of new variants of coronaviruses and a scale of total infected people is closely related to the probability *k*. The typical value of *k* in Japan evaluated in this study is smaller than those in European countries for the *δ* surge, but it gets close to European ones for the *o* surge.

## Data Availability

All data produced in the present work are contained in the manuscript.

## Author Contributions

Y.I. developed and formulated the model. K.S. and T.N. contributed to the data analysis. All authors contributed to the interpretation of the results obtained in this study and the final manuscript.

## Institutional Review Board Statement

Not applicable

## Informed Consent Statement

Not applicable

## Data Availability Statement

Not applicable

## Acknowledgments

We thank Prof. Yoshiharu Matsuura, Prof. Fumio Ohtake and all the member of Division of Scientific Information and Public Policy (SiPP) at Center for Infectious Disease Education and Research (CiDER) Osaka University for useful discussions and comments. We also thank Mr. Toru Ohmuta for the comments.

